# Genetic analysis of *VCP* variants in a Turkish dementia cohort

**DOI:** 10.1101/2023.02.13.23285395

**Authors:** Nadia Dehghani, Zeynep Tufekcioglu, Gamze Guven, Kaitlyn Westra, Hasmet Hanagasi, Ebba Lohmann, Bedia Samanci, Hakan Gurvit, Basar Bilgic, Rita Guerreiro, Murat Emre, Jose Bras

**Affiliations:** Department of Neurodegenerative Science, Van Andel Research Institute, Grand Rapids, MI, USA; Department of Neurology, Faculty of Medicine, Istanbul Aydin University, Istanbul, Turkey; Department of Genetics, Aziz Sancar Institute of Experimental Medicine, Istanbul University, Istanbul, Turkey; Behavioural Neurology and Movement Disorders Unit, Department of Neurology, Istanbul Faculty of Medicine, Istanbul University, Istanbul, Turkey; Department of Neurodegenerative Diseases, Hertie Institute for Clinical Brain Research, University of Tübingen, Tübingen, Germany; DZNE, German Center for Neurodegenerative Diseases, Tübingen, Germany; Division of Psychiatry and Behavioral Medicine, Michigan State University College of Human Medicine, Grand Rapids, MI, USA

**Author notes:** **Correspondence:** Jose Bras.

**Keywords:** Dementia, VCP, DLB, Exomes, Turkey

## Abstract

*Valosin-containing protein* (*VCP*) mutations are causative for multisystem proteinopathy, a disease characterized by variable phenotypes including inclusion body myopathy, Paget’s disease of bone, and frontotemporal dementia. More recent reports identified *VCP* variants as the cause of other neurodegenerative diseases, such as Parkinson’s disease and vacuolar tauopathy. We screened a Turkish dementia cohort for *VCP* variants in order to assess their role as the cause of disease in this population. One hundred and forty six Turkish dementia patients were examined clinically and were analyzed for *VCP* coding variants using whole-exome sequencing. Familial samples were collected and analyzed in order to test for segregation of candidate variants. We identified a heterozygous missense VCP p.Ile216Met variant segregating with disease in a family where the proband was diagnosed with prodromal dementia with Lewy bodies. Our report potentially extends the spectrum of phenotypes attributed to *VCP* mutations to include prodromal dementia with Lewy bodies.

## Introduction

Valosin-containing protein (VCP) belongs to the AAA+ ATPase family and is involved in many molecular pathways, particularly concerning protein degradation such as the ubiquitin-proteasome and lysosomal systems (Meyer et al., 2012). VCP contains an N-domain and a C-terminus surrounding two ATPase domains (D1 and D2). Heterozygous mutations in *VCP* are causative for multisystem proteinopathy (MSP) which can present with a variety of phenotypes including inclusion body myopathy, Paget’s disease of bone, and frontotemporal dementia (IBMPFD) (**Figure 1A**). Such mutations, mostly located within the N-domain and ATPase domains, have been reported to increase ATPase activity (Niwa et al., 2012). Recent work further characterizing the effects of *VCP* variants in MSP, reported that they result in faster substrate binding and unfolding when specifically studying the interaction between VCP and the VCP substrates NPLOC4 and UFD1L (Blythe et al., 2017, 2019).

**Figure 1.**
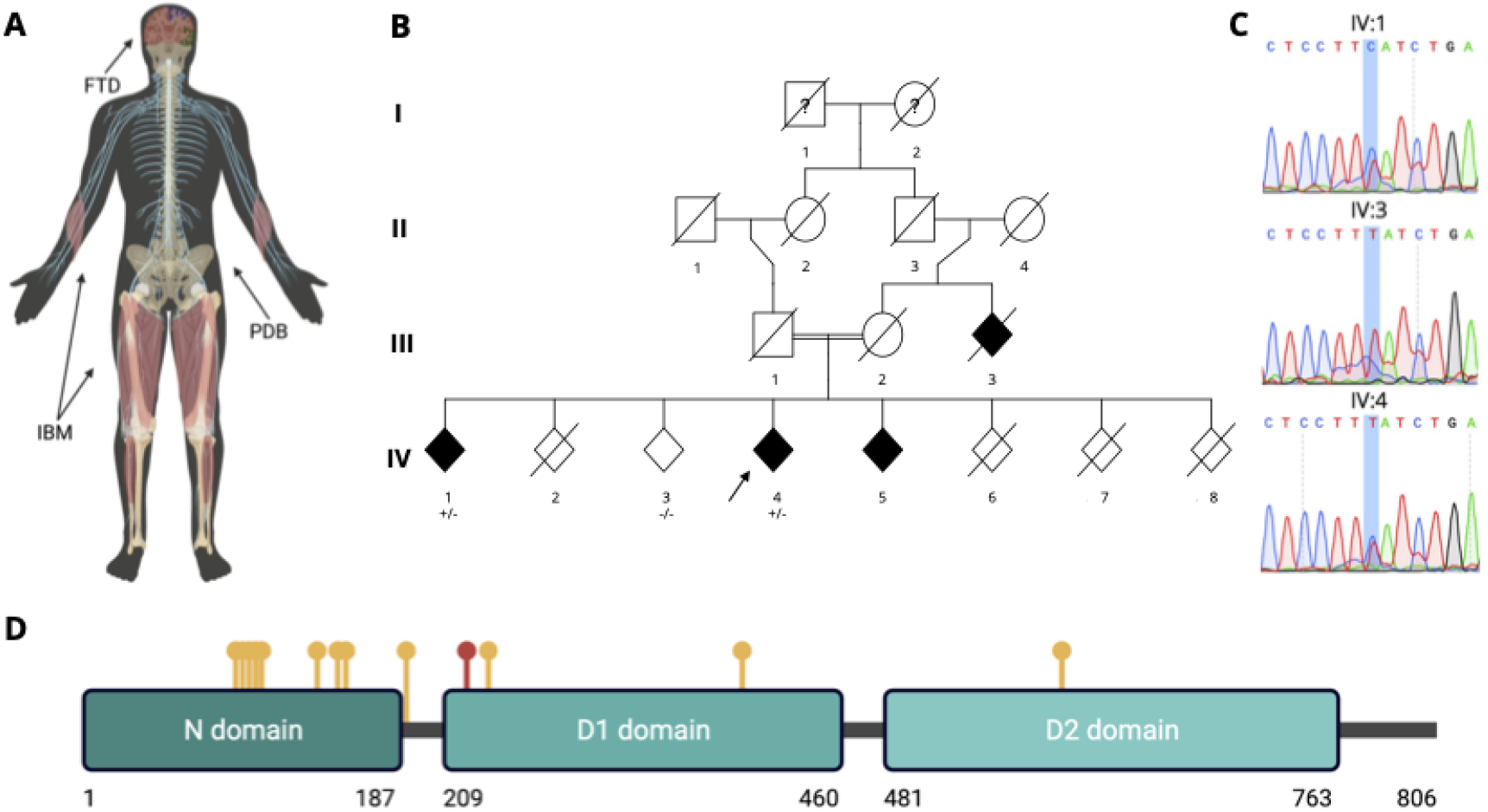
**A:** *A schematic of the phenotypes presenting in multisystem proteinopathy*. Phenotypes encompass inclusion body myopathy, Paget’s disease of bone, and frontotemporal dementia. Arrows point to body regions that are commonly affected by each phenotype. **B:** *Partial pedigree tree for the Turkish pDLB patient with VCP p.Ile216Met variant*. Square = male; circle = female; diamond = unknown sex; black = individuals affected with dementia; white = individuals with no reported dementia phenotype; diagonal line = deceased individual; double bar = consanguineous marriage; arrow = the proband. Where tested, “+/-” indicates a heterozygous variant carrier and “-/-” indicates an individual not carrying the variant of interest. An affected sibling (IV:1) previously diagnosed with AD had an age of onset in their mid-70s, and a 5-year disease duration. They are currently entirely dependent. An unaffected sibling (IV:2) died from non-dementia reasons. Another unaffected sibling (IV:3) in their early 70s had DNA available and Sanger sequencing was performed to test for this variant. The proband (IV:4) had an age of onset in their early 60s. An affected sibling (IV:5) previously diagnosed with AD had an age of onset in their mid-50s, and 5-year disease duration. They are now fully dependent. It was not possible to obtain a sample from this sibling. A sibling with Down’s syndrome (IV:6) died when they were in their late teens. The unaffected sibling (IV:7) died in their late teens because of an accident. Another unaffected sibling (IV:8) died in their 50s due to non-dementia related reasons. **C:** *Sanger traces for proband and two siblings*. These show the heterozygous variant present in both the proband (IV:4) and affected sibling (IV:1), with no variant in the unaffected sibling (IV:3). **D:** *VCP protein domains*. Domain positions are as reported by DeLaBarre and Brunger, 2003. VCP p.Ile216Met is marked in red and previously reported variants are marked in yellow (see **Supplementary Table 4** for genomic positions and a description of the phenotypes reported for variant carriers). Panels A and D were created with BioRender.com.

In addition to MSP, *VCP* mutations have also been reported to cause autosomal dominant Charcot-Marie Tooth disease type 2Y (Gonzalez et al., 2014; Jerath et al., 2015) and amyotrophic lateral sclerosis with or without frontotemporal dementia (FTD) (Johnson et al., 2010). *VCP* has also been investigated in Parkinson’s disease (PD) (Majounie et al., 2012; Regensburger et al., 2017) with decreased *VCP* expression suggested as a biomarker for early-onset PD (Alieva et al., 2020). *VCP* variants have also been reported in Alzheimer’s disease (AD); one study specifically screened *VCP* following linkage near this locus on chromosome 9 and identified VCP:p.Arg92His in 2% of the AD siblings contributing to the linkage peak, and rs10972300-T which demonstrated an association with AD in the US cases but not those from the UK with no further reports of replication (Kaleem et al., 2007; Mehta et al., 2013). Moreover, in one family with VCP:p.Arg155His, amyotrophic lateral sclerosis was reported in two individuals, and neuropathological-confirmed AD was reported in one individual (Kaleem et al., 2007; Mehta et al., 2013). Most recently, VCP p.Asp395Gly was reported in 2 families from the US and Greece with vacuolar tauopathy, a form of FTD characterized by neuronal vacuoles and tau neurofibrillary tangles; the authors suggested that VCP functions as a tau disaggregase (Darwich et al., 2020).

As the number of diseases and populations studied for *VCP* variants increases, we set out to specifically investigate *VCP* variants in a Turkish dementia cohort.

## Materials and Methods

### Ethics Approval and Consent to Participate

This study was approved by the Ethics Committee of Istanbul Faculty of Medicine, Istanbul University. A neurologist took the necessary clinical information after obtaining informed written consent for study participation from the patients and their participating family members. Consent was provided by the legally authorized representative for subjects unable to consent. Research was performed in accordance with the Declaration of Helsinki.

### Participants

One hundred and forty six probands were whole-exome sequenced (**Table 1**); 19 of these carry previously published definite or potentially pathogenic variants in the genes *PSEN1, PSEN2, MAPT, GRN, TREM2, NOTCH3* and *C9orf72* (**Supplementary Table 1**). The dementia phenotype includes individuals with an unclear or atypical diagnosis.

**Table 1.** Characteristics for the 146 Turkish dementia patients in this study. AD - Alzheimer’s disease, FTD - frontotemporal dementia, MCI - mild cognitive impairment, LBD - Lewy body dementia (includes both dementia with Lewy bodies (DLB) and Parkinson’s disease dementia (PDD)), CADASIL - cerebral autosomal dominant arteriopathy with subcortical infarcts and leukoencephalopathy, PCA - posterior cortical atrophy, CBD - corticobasal degeneration.

| Clinical Phenotype | Sex # Male \| # Female (Total) | Mean AAO (±SD) |
| --- | --- | --- |
| AD | 27\|42 (69) | 64.4±12.4 |
| FTD | 24\|17 (41) | 56.7±14.5 |
| MCI | 4\|7 (11) | 66.2±8.4 |
| LBD | 5\|1 (6) | 62.0±5.8 |
| CADASIL | 1\|1 (2) | 55.0±12.7 |
| PCA | 1\|1 (2) | 42.0±5.7 |
| CBD | 0\|1 (1) | Early 50s |
| Unspecified dementia | 10\|4 (14) | 58.9±15.9 |

All participants were recruited at the Behavioral Neurology and Movement Disorders Unit outpatient clinic of Istanbul Faculty of Medicine, Istanbul University. They underwent detailed clinical and neuropsychological examination, brain magnetic resonance imaging (if this was not possible, computerized tomography was performed), and FDG-PET or CSF biomarkers when necessary. Depending on the admission date of the patients, diagnosis of Alzheimer’s dementia was based on the National Institute of Neurological and Communicative Disorders and Stroke and the Alzheimer’s Disease and Related Disorders Association (NINCDS/ADRDA) (McKhann et al., 1984; McKhann et al., 2011). Mild cognitive impairment (MCI) patients were diagnosed as described by Frank and Petersen (Frank and Petersen, 2008) and Albert et al. (Albert et al., 2011). FTD was defined following the Lund and Manchester criteria (The Lund and Manchester Groups, 1994) and as described by Rascovsky et al. (Rascovsky et al., 2011). The criteria defined by Gorno-Tempini et al. was used for classification of primary progressive aphasia and its variants (Gorno-Tempini et al., 2011). The consortium on DLB international workshop criteria was used for DLB diagnosis (McKeith et al., 1996; McKeith et al., 2017). Prodromal dementia with Lewy bodies (pDLB) was diagnosed according to the McKeith criteria (McKeith et al., 2020). Crutch et al. was used for the diagnosis of posterior cortical atrophy (Crutch et al., 2017). Armstrong et al. was used for the diagnosis of corticobasal degeneration (Armstrong et al., 2013). Emre et al. was used for the diagnosis of Parkinson’s disease dementia (Emre et al., 2007). The diagnosis of CADASIL was based on supportive clinical and neuroimaging features and molecular genetic analysis results.

### Exome Sequencing

Genomic DNA was extracted from peripheral blood using the Roche MagNA Pure DNA isolation kit. Exome sequencing libraries were prepared using the SureSelect Exome Capture Kit v4, 6 or 7 (Agilent), TruSeq Exome Capture Kit, or SeqCap EZ Exome Library v1. Sequencing was performed on Illumina HiSeq2000, NovaSeq6000, NextSeq500, or NextSeq550 with 75-150 bp paired-end reads. Coverage of at least 30x was achieved and exome data processing was accomplished following the GenomeAnalysisTK (GATK) best practices v4 (McKenna et al., 2010). In short, alignment was performed with the Burrows-Wheeler Aligner (bwa-mem) v0.7.17 against the hg38 genome assembly, samblaster v0.1.24 identified duplicates and base recalibration was achieved with GATK v4.1.4.1, excluding known sites. Variant Quality Score Recalibration (VQSR) was applied to variants which were excluded if they did not reach a 0.99 score (DePristo et al., 2011; Van der Auwera et al., 2013). Finally, data was annotated with snpEff v4.3 (Cingolani et al., 2012), dbNSFP v4.0 (Liu et al., 2013) and dbSNP v151 (Sherry et al., 2001).

### Genetic Quality Control and Filtering

We searched for *VCP* coding variants and filtered these for genotyping quality ≥99 and minor allele ratio between 0.3-0.75 for heterozygous variants, and ≥0.9 for homozygous variants. In the event that *VCP* coding variants passed our variant filters, we further screened the same patient data for exonic variants (with total AF ≤0.01 in the genome aggregation database (gnomAD) v3 (Karczewski et al., 2020)) in genes known to cause, or be associated with risk of neurodegenerative diseases (**Supplementary Table 2**). We also considered the allele frequency from the Greater Middle East (GME) Variome (Scott et al., 2016) and the Turkish (TR) Variome (Kars et al., 2021).

### Sanger Sequencing

The region encompassing the *VCP* variant NM_007126.5:c.648A>G was polymerase chain reaction (PCR) amplified using Roche FastStart PCR Master Mix polymerase (Roche Diagnostics, Corp., Indianapolis, IN, USA) and the following primers: 5′-GCCTTCTCCTTTCTTTATCTCCTCC-3′ and 5′-CATTGGCACCACTTTAGACTTGA-3′. Each PCR product was sequenced using the same forward and reverse primers with Applied Biosystems BigDye version 3.1 and run on an ABI3730xl (Applied Biosystems, Carlsbad, CA, USA) genetic analyzer as per manufacturer’s instructions. The sequences were analyzed with SnapGene software, version 4.3.7 (from Insightful Science; available at snapgene.com).

### Additional datasets

We screened both the UK Biobank Research Analysis Platform and additional dementia samples (including genomes from 2610 Lewy body dementia, 2242 frontotemporal dementia and 1920 controls [dbGaP Study Accession: phs001963.v1.p1] and 1118 DLB exomes (Orme et al., 2020) for carriers of candidate variants.

## Results

We identified a *VCP* coding variant in one individual within the studied Turkish dementia cohort. This was a heterozygous missense *VCP* variant NM_007126.5:c.648A>G in an individual diagnosed with prodromal dementia with Lewy bodies (pDLB) according to the McKeith criteria (McKeith et al., 2020). This equates to VCP p.Ile216Met at the protein level.

This patient in their mid-60s was first assessed in their early 60s for cognitive decline. They were reported to have cognitive problems two years before initial assessment. The main complaints from both the patient and their family regarded cognitive slowness and perception problems. They had no psychotic or other behavioral symptoms. In cognitive testing their mini-mental state examination score was 29/30, their main cognitive deficit was reduced verbal fluency. They reported multiple autonomic symptoms including urinary incontinence, constipation, orthostatic hypotension, and rapid eye movement sleep behavior disorder. The clinical diagnosis was pDLB in accordance with diagnostic criteria for this disorder (McKeith et al., 2020). Based on whole-exome sequencing data, this proband is homozygous for the APOE e3 allele.

Their family history was remarkable for dementia (**Figure 1B**). The proband’s relative, now in their early 90s, (individual III:3 in the pedigree) had dementia for three years before death (age of onset in their mid-80s, they died of cancer after five years). The patient had consanguineous parents (aunt-uncle children). The proband’s mother died when she was in her late 80s and was reported to be unaffected at the time of death. The proband’s father died when he was in his mid-70s without dementia. The mother also had consanguineous parents.

To test for segregation, we Sanger-sequenced DNA available from the proband and two siblings. We confirmed the presence of the variant in the proband and observed the same variant in the affected sibling (individual IV:1 in the pedigree) and no variant in the unaffected sibling (individual IV:3 in the pedigree) (**Figure 1C**).

VCP p.Ile216Met is located within the D1 ATPase domain (**Figure 1D**) and VCP p.Ile216 is well-conserved across different species, suggesting functional importance of variants at this position (see **Supplementary Figure 1** for multiple sequence alignment).. The variant was considered damaging and deleterious by *in silico* prediction tools including PolyPhen-2, SIFT, and CADD (Kumar et al., 2009; Adzhubei et al., 2010; Kircher et al., 2014). Moreover, it was not reported in either gnomAD, the GME Variome, or the TR Variome. There were also no additional candidate variants (and specifically no homozygous variants) identified in this case when exonic variants in genes known to cause, or be associated with risk of neurodegenerative diseases were screened (see **Supplementary Table 2** for this gene list and **Supplementary Table 3** for the variants identified in our proband). This suggests that this variant is both rare and is not a common population-specific variant. This proband also does not carry previously published definite or potentially pathogenic variants as summarized in **Supplementary Table 1**. Furthermore, we did not identify other VCP Ile216Met carriers when we consulted additional datasets (see Methods) or any VCP variant carriers with similar phenotypes when we consulted GeneMatcher (Sobreira et al., 2015).

## Discussion

Here we report a heterozygous *VCP* variant (NM_007126.5:c.648A>G; p.Ile216Met) in a pDLB patient. This missense variant has been previously reported on ClinVar with uncertain significance for IBMPFD (submitted by Invitae).

Although the absence of dementia in the proband’s parents (Individuals III:1 and III:2 in the pedigree) and the multiple loops of consanguinity in the family would suggest an autosomal recessive mode of disease inheritance, there are several reasons why a heterozygous variant could be the cause of disease in this family. The variant might present with incomplete penetrance, or either parent might have developed the disease at a later age. In particular, the father can be considered as a potential carrier based on the observed consanguinity and age at death identical to age at disease onset of sibling IV:1. Moreover, it should be noted that the maternal relative was diagnosed with dementia when in their mid-80s. The clinical picture described in this family carrying the VCP p.Ile216Met variant does not represent a known clinical phenotype for pathogenic *VCP* mutations (**Supplementary Table 4**); however, some similarities between the proband described here and previous reports of *VCP* variant carriers with PD include REM sleep behavior disorder, urinary incontinence, and absence of psychosis (Chan et al., 2012; Spina et al., 2013; Regensburger et al., 2017). Sphincter disturbances were also reported in a large MSP pedigree (Miller et al., 2009). To date, no correlations between mutations and the incidence of specific clinical phenotypes have been described, with the exception of the association of p.R159C with a later age of onset of myopathy and absence of Paget’s disease of bone when compared to other mutations (Al-Obeidi et al., 2018).

A limitation of this current study is the lack of certainty regarding the pathogenicity of VCP p.Ile216Met. Based on the data available, it is not possible to definitively conclude that this is the Mendelian cause of disease. Still, the association of this variant with the disease is supported by several lines of evidence: 1) the limited (due to sample availability) segregation of the variant with disease in the family; 2) the very low population frequency of the variant, being absent from population databases and only reported once in ClinVar; and 3) by similarities with clinical features described for other *VCP* variant carriers.

If this VCP variant is indeed the cause of disease in the studied family, it may have an effect on disease independently or through the development of Lewy body pathology. Given that VCP immunoreactivity has been shown in Lewy bodies of cases with DLB (Mori et al., 2013), one would expect the latter to be more probable, however, we cannot conclude that this patient has Lewy pathology without autopsy confirmation.

Considering the previous genotype-phenotype correlations for *VCP* mutations, one might have expected for this *VCP* variant to present as FTD. In this respect, it is interesting to note that a clinical overlap between FTD and DLB has been reported with some patients described as presenting clinical features suggestive of both FTD and DLB (Claassen et al., 2008). The clinical picture for *GRN* mutation carriers is also broad, with reports including, among others, DLB and AD (Orme et al., 2020); this is intriguing since *GRN* variants are a known Mendelian cause of FTD.

DLB is a relatively understudied clinical phenotype accounting for at least 7.5% of all dementias in clinic-based studies (Walker et al., 2015). The potential under- and misdiagnosis of DLB is reinforced by the knowledge that the two affected siblings (one of which had a sample available and was determined to also carry the same *VCP* variant) had both previously received a diagnosis of Alzheimer’s disease. On the other hand, perhaps this also suggests a pathological mechanism, induced by the *VCP* variant, that causes protein aggregation and leads to dementia, independently of Lewy body formation.

To the best of our knowledge, this is the first report of a *VCP* variant in pDLB, and the first report of a potentially pathogenic *VCP* variant in a Turkish family. Although we have not been able to identify a second DLB case carrying this same variant, it is still important to describe these results to help corroborate potentially similar future findings.

## Data Availability

Data generated during this study is included in this published article and supplementary information files. We have submitted this VCP variant (NM_007126.5:c.648A>G) to ClinVar (accession number: SCV002524135).

## Acknowledgements

This research has been conducted using data from UK Biobank, a major biomedical database (www.ukbiobank.ac.uk).

The authors acknowledge the contribution of data from DEMENTIA-SEQ: Genome sequencing in Lewy Body Dementia, Frontotemporal Dementia, and neurologically healthy controls, accessed through the database of Genotypes and Phenotypes (phs001963.v1.p1).

## Author Contributions

ND, RG and JB designed the study and wrote the manuscript. ND, KW, RG and JB performed data analyses and interpreted the data. ZT, GG, HH, EL, BS, HG, BB and ME collected and characterized samples for inclusion in the study. All authors provided critical feedback and helped shape the research, analysis and manuscript. All authors read and approved the final manuscript.

## Data Availability Statement

Data generated during this study is included in this published article and supplementary information files. We have submitted this *VCP* variant (NM_007126.5:c.648A>G) to ClinVar (accession number: SCV002524135).

