## Supplementary Material for "Genetic analysis of *VCP* variants in a Turkish dementia cohort"

| Variant | HGVS | Number of individuals from our cohort | Phenotype | Reference |
|---|---|---|---|---|
| TREM2 p.Tyr38Cys | ENST00000373113.3:c.113A>G | 1 | FTD | [1] |
| TREM2 p.Thr66Met | ENST00000373113.3:c.197C>T | 1 | FTD | [1] |
| TREM2 p.Gln33Ter | ENST00000373113:c.97C>T | 1 | FTD | [1] |
| PSEN1 p.Pro264Leu | ENST00000324501:c.791C>T | 1 | FTD | [2] |
| PSEN1 p.His214Tyr | ENST00000324501:c.640C>T | 1 | FTD | [2] |
| PSEN1 p.His163Arg | ENST00000324501:c.488A>G | 1 | AD | [2] |
| PSEN1 p.Leu134Arg | ENST00000324501:c.401T>G | 1 | dementia | [2] |
| PSEN1 p.Leu262Val | ENST00000324501:c.784T>G | 1 | AD | [2] |
| PSEN2 p.Ser130Leu | ENST00000366783.3:c.389C>T | 1 | AD | [2] |
| PSEN2 p.Met174Val | ENST00000366783.3:c.520A>G | 2 | 1 FTD, 1 dementia | [2] |
| C9orf72 expansion | ENST00000379997.3: c.-45+(GGGGCC)$^n$ | 3 | 3 FTD | [3] |
| NOTCH3 p.Arg1231Cys | ENST00000263388.2:c.3691C>T | 1 | AD | [4] |
| MAPT p.Pro636Leu | ENST00000344290.5:c.1907C>T | 1 | FTD | [3] |
| GRN p.Cys139Arg + c.-22C>T | ENST00000053867.3:c.415T>C + ENST00000053867.3:c.-22C>T | 1 | FTD | [3] |
| GRN c.708+6_9delTGAG | ENST00000053867:c.708+6_708+9delTGAG | 1 | AD | [3] |
| GRN p.Pro209Leu | ENST00000053867:c.626C>T | 1 | PCA | [3] |

**Supplementary Table 1.** Previously published definite or potentially pathogenic variants in this dementia cohort.

| | | | | | | |
|---|---|---|---|---|---|---|
| **AD** | | | | | | |
| *APP* | *PSEN1* | *PSEN2* | *APOE* | *TREM2* | *CLU* | *PICALM* |
| *CR1* | *BIN1* | *MS4A6A* | *MS4A4E* | *CD33* | *ABCA7* | *CD2AP* |
| *EPHA1* | *HLA-DRB5* | *HLA-DRB1* | *SORL1* | *PTK2B* | *SLC24A4* | *ZCWPW1* |
| *CELF1* | *FERMT2* | *CASS4* | *INPP5D* | *MEF2C* | *NME8* | |
| **PD** | | | | | | |
| *SNCA* | *PARK2/PRKN* | *PINK1* | *PARK7/DJ-1* | *LRRK2* | *PLA2G6* | *FBXO7* |
| *VPS35* | *ATP13A2* | *DNAJC6* | *SYNJ1* | *GBA* | *MAPT* | *RAB7L1* |
| *BST1* | *GAK* | *ACMSD* | *STK39* | *SYT11* | *FGF20* | *STX1B* |
| *GPNMB* | *SIPA1L2* | *INPP5F* | *MIR4697HG* | *GCH1* | *VPS13C* | *DDRGK1* |
| *MCCC1* | *SCARB2* | *CCDC62* | *RIT2* | *SREBF1* | | |
| **FTD** | | | | | | |
| *GRN* | *CHMP2B* | *HNRNPA1* | *HNRNPA2B1* | *SQSTM1* | *OPTN* | *CHCHD10* |
| *VCP* | *SIGMAR1* | *PRKAR1B* | *TMEM106B* | *UBQLN2* | *ATXN2* | |
| **DLB** | | | | | | |
| *CNTN1* | *CYP2D6* | *EIF4G1* | *BCL7C/STX1B* | *GABRB3* | *GIGYF2* | *PRNP* |
| *SNCB* | *SOX17* | *GBA* | *APOE* | *SNCA* | | |
| **Additional genes** | | | | | | |
| *ANG* | *CCNF* | *CSF1R* | *CTSC* | *CYLD* | *DCTN1* | *FUS* |
| *GLE1* | *ITM2B* | *MATR3* | *NEK1* | *NOTCH3* | *PFN1* | *PNPLA6* |
| *RAB38* | *SERPINI1* | *SOD1* | *TAF15* | *TARDBP* | *TBK1* | *TIA1* |
| *TUBA4A* | *TYROBP* | *VAPB* | | | | |

**Supplementary Table 2.** List of genes known to cause, or be associated with risk of neurodegenerative diseases, as previously described [5].

| Gene | Variant | Position, hg38 (position in hg19) | Reason for excluding | Total allele frequency in gnomAD v3.1.1 (hg38) | Total allele frequency in gnomAD v2.1.1 (hg19) | Frequency in GME Variome (hg19) | Frequency in the TR Variome |
|---|---|---|---|---|---|---|---|
| *OPTN* | p.His29Pro (c.86A>C) | Chr10:13109208 (chr10:13151208) | Previously reported pathogenic variants in this region are frameshifts and stop-gains | Not reported | Not reported | Not reported | Not reported |
| *SORL1* | p.Cys1271Cys (c.3813C>T) | Chr11:121586328 (chr11:121457037) | Synonymous | 1.97e-5 | 4.38e-5 | Not reported | 0.000744934 (5 heterozygous carriers) |
| *TUBA4A* | p.Arg123Cys (c.367C>T) | Chr2:219251573 (chr2:220116295) | Reports in databases | Not reported | 1.99e-5 | 0.0005 ( 1 heterozygous individual from Northeast Africa) | Not reported |
| *VCP* | p.Ile216Met (c.648A>G) | Chr9:35064214 (chr9:35064211) | NA- candidate variant | Not reported | Not reported | Not reported | Not reported |
| *ZCWPW1* | p.Glu105Gly (c.314A>G) | Chr7:100419158 (chr7:100016781) | Reports in databases | 4.18e-3 | 5.74e-3 | 0.0232 (includes 10 heterozygous individuals from the Turkish Peninsula) | 0.025610482 (7 homozygous carriers, 158 heterozygous carriers) |

**Supplementary Table 3.** Exonic variants in the proband in genes known to cause, or be associated with risk of neurodegenerative diseases. These were either absent or reported in gnomAD v3 with a total AF ≤0.01. All of these variants were heterozygous in the proband.

| **Variant** | **Phenotype (Reference or ClinVar submitter)** |
|---|---|
| NM_007126.5(**VCP**):c.1774G>A (p.Asp592Asn) | ALS [6]. |
| NM_007126.5(**VCP**):c.1184A>G (p.Asp395Gly) | Vacuolar tauopathy [7]. |
| NM_007126.5(**VCP**):c.695C>A (p.Ala232Glu) | Myopathy, fractures and PDB [8]. |
| NM_007126.5(**VCP**):c.572G>C (p.Arg191Pro) | ALS with or without FTD (Suna and Inan Kirac Foundation Neurodegeneration Research Laboratory, Koc University). |
| NM_007126.5(**VCP**):c.572G>A (p.Arg191Gln) | IBMPFD and IBM only [9], limb girdle muscular dystrophy [10], muscle weakness, Paget's disease of bone and cognitive impairment [11], IBMPFD with speech, memory and emotional problems [12], muscle weakness and wasting, and Parkinsonism [13], clinical phenotype resembling facioscapulohumeral muscular dystrophy (one patient showed transversal smile and difficulty to blow, along with mild dysexecutive syndrome); neither patient showed sign of Paget's disease or rimmed vacuoles upon muscle biopsy which would indicate inclusion body myopathy [14], ALS in an Italian kindred [6], family from the US or Canada with mutation carrier presenting IBM; additional affected family members also presented with PDB and FTD [8], IBM with early-onset PDB and FTD (Center of Genomic medicine, Geneva,University Hospital of Geneva), ALS with or without FTD (Institute of Human Genetics,Cologne University). |
| NM_007126.5(**VCP**):c.476G>A (p.Arg159His) | A Belgian MSP patient homozygous for this variant presented with earlier age at onset [15], reported in 3 Belgian FTD patients [16], a Belgian man with axonal sensory neuropathy, FTD and asymptomatic PDB [17], one familial ALS patient from The Netherlands [18], 2 Belgian families presenting with FTD, PDB or both, and one Austrian family with IBM, PDB and dementia [19, 20], IBMPFD without PDB or cognitive symptoms [21]. |
| NM_007126.5(**VCP**):c.475C>A (p.Arg159Ser) | Dutch FTD patient [22]. |
| NM_007126.5(**VCP**):c.475C>T (p.Arg159Cys) | ALS with or without FTD (Suna and Inan Kirac Foundation Neurodegeneration Research Laboratory, Koc University), 2 families with IBM with or without FTD, with two individuals from the same family diagnosed with PD [9], one family diagnosed with ALS without dementia or muscle disease; one affected member was also diagnosed with PDB before onset of neuromuscular symptoms; this variant was also present in an unaffected family member older than the latest age at onset reported to date [23], an American man with likely German descent diagnosed with PD with symptoms including likely REM sleep behavior disorder and family history of FTD and IBM [24], an Italian man with IBM and FTD [25], female |

| | |
|---|---|
| | with no evident muscle weakness or neuropathy, no PDB at age 59 years [26]. |
| NM_007126.5(**VCP**):c.475C>G (p.Arg159Gly) | A US family presenting with ALS and either dementia or PDB [6]. |
| NM_007126.5(**VCP**):c.464G>C (p.Arg155Pro) | 2 families presenting IBM and/or PDB, or IBM and FTD [9], identified in the proband of 1 family from the US or Canada presenting IBM and PDB [8]. |
| NM_007126.5(**VCP**):c.464G>A (p.Arg155His) | Seven families from the US or Canada with the mutation carrier presenting IBM, PDB, or both, or asymptomatic to date with a family history of IBMPFD [8], IBM with early-onset PDB with or without FTD (Equipe Genetique des Anomalies du Developpement, Université de Bourgogne), IBMPFD with episodic memory problems and increased irritability [12], 6 patients presenting with IBM and/or PDB, one of which presenting with IBM and ALS [27], pathologically confirmed ALS without evidence of IBM or PDB [6], male with muscle weakness, PDB and no cognitive impairment [26], 6 patients with IBM and/or PDB, and/or FTD [28], 4 Italian family members presenting with IBM and FTD with or without PDB; two of whom presented with myopathic changes and depression [29], 1 patient with progressive muscle weakness and atrophy, PDB confined to the first lumbar vertebra, and mild FTD following neuropsychological examination [30]. |
| NM_007126.5(**VCP**):c.463C>T (p.Arg155Cys) | ALS with or without FTD (Suna and Inan Kirac Foundation Neurodegeneration Research Laboratory, Koc University), 11 UK families with a combination of myopathic changes, PDB, cognitive impairment, sphincter involvement and abnormal echocardiogram [11], earlier age at onset of myopathy and PDB, and reduced mean survival, compared to p.Arg155His; one family was also diagnosed with ALS [31], 3 siblings from a Korean family presenting with FTD and PDB, IBM, or both [32], 5 IBMPFD patients from France or Spain all first presenting with myopathy, and 2 with dysexecutive syndrome [33], an Italian family with progressive myopathy and dementia or preclinical signs of PDB [34], female with progressive proximal muscle weakness, FTD and severe generalized wasting of skeletal muscles with no signs of PDB [30], a family presenting with proximal and axial muscle weakness with myotonia and/or FTD [35], two families from the US or Canada with mutation carriers presenting IBM with or without PDB [8], IBMPFD patient [12], one patient with IBM and FTD [28]. |
| NM_007126.5(**VCP**):c.409C>T (p.Pro137Ser) | Identified in a patient referred with suspected prion disease who also carries a de novo PRNP variant [36]. |
| NM_007126.5(**VCP**):c.294T>A (p.Asp98Glu) | IBM with early-onset PDB with or without FTD (Institute of Human Genetics, University of Leipzig Medical Center). |
| NM_007126.5(**VCP**):c.290G>A (p.Gly97Glu) | 5 individuals from the same Chinese family carried this variant and presented with IBM; 3 had PDB and none had FTD although 2 were |

| | |
|---|---|
| | diagnosed with PD [37], an individual of Dutch and Italian descent presenting with a lower motor neuron syndrome and axonal neuropathy [38]. |
| NM_007126.5(**VCP**):c.283C>T (p.Arg95Cys) | ALS (Suna and Inan Kirac Foundation Neurodegeneration Research Laboratory, Koc University), spastic paraplegia (Paris Brain Institute,Inserm - ICM), sporadic IBM [39]. |
| NM_007126.5(**VCP**):c.283C>G (p.Arg95Gly) | Identified in a proband with family history of IBM, with or without FTD; their sibling also presents with PDB only [8]. |
| NM_007126.5(**VCP**):c.277C>T (p.Arg93Cys) | Reported in a German distal myopathy patient [40], two unrelated individuals in the UK presenting with cognitive impairment and sphincter involvement in one [11], a family presenting with FTD together with either IBM, PDB, or both [35]. |
| NM_007126.5(**VCP**):c.271A>T (p.Asn91Tyr) | A Brazilian family with individuals presenting with either IBM, ALS or FTD (no PDB) [41]. |

**Supplementary Table 4.** Phenotypic information for missense *VCP* variants reported as pathogenic/likely pathogenic on ClinVar with accompanying citation or reported condition. ALS, amyotrophic lateral sclerosis; FTD, frontotemporal dementia; IBM, inclusion body myopathy; IBMPFD, inclusion body myopathy, Paget's disease of bone, frontotemporal dementia; PD, Parkinson's disease.

**Supplementary Figure 1.** Multiple sequence alignment for VCP across different species. *=conserved sequence, :=conservative mutation, .=semi-conservative mutation. Performed with Clustal Omega [42–44]. The conservation of amino acid 216 in Homo sapiens is highlighted in red.

```
CLUSTAL O(1.2.4) multiple sequence alignment

AIG99085.1[Archaeoglobus_fulgidus_DSM_8774]        QAVVFVAVKTEPKGPVVIDETTKVVYRDRPAKGF-ERFGKAGITYEDIGGLKEELQKVRE  211
NP_010157.1[Saccharomyces_cerevisiae_S288C]        RQVEFKVVDVEPEEYAVVAQDTIIHWEGEPINREDEENNMNEVGYDDIGGCRKQMAQIRE  228
NP_477369.1[Drosophila_melanogaster]               RPIEFKVVLTDPEPYCIVAPETVIFCDGDPIKREEEEESLNAVGYDDIGGCRKQLAQIKE  215
NP_958889.1[Danio_rerio]                           RAVEFKVVETDPSPYCIVAPDTVIHCEGEPIKREDEEESLNEVGYDDIGGVRKQLAQIKE  218
XP_032069281.1[Thamnophis_elegans]                 RAVEFKVVETDPSPYCIVAPDTVIHCEGEPIKREDEEESLNEVGYDDIGGCRKQLAQIKE  218
NP_001029466.1[Bos_taurus]                         RAVEFKVVETDPSPYCIVAPDTVIHCEGEPIKREDEEESLNEVGYDDIGGCRKQLAQIKE  218
XP_038537953.1[Canis_lupus_familiaris]             RAVEFKVVETDPSPYCIVAPDTVIHCEGEPIKREDEEESLNEVGYDDIGGCRKQLAQIKE  218
NP_446316.1[Rattus_norvegicus]                     RAVEFKVVETDPSPYCIVAPDTVIHCEGEPIKREDEEESLNEVGYDDIGGCRKQLAQIKE  218
NP_009057.1[Homo_sapiens]                          RAVEFKVVETDPSPYCIVAPDTVIHCEGEPIKREDEEESLNEVGYDDIGGCRKQLAQIKE  218
XP_014972997.1[Macaca_mulatta]                     RAVEFKVVETDPSPYCIVAPDTVIHCEGEPIKREDEEESLNEVGYDDIGGCRKQLAQIKE  218
NP_033529.3[Mus_musculus]                          RAVEFKVVETDPSPYCIVAPDTVIHCEGEPIKREDEEESLNEVGYDDIGGCRKQLAQIKE  218
NP_001267410.1[Pan_troglodytes]                    RAVEFKVVETDHSPYCIVAPDTVIHCEGEPIKREDEEESLNEVGYDDIGGCRKQLAQIKE  218
NP_001095217.1[Xenopus_laevis]                     RAVEFKVVETDPSPYCIVAPDTVIHCEGEPIKREDEEESLNEVGYDDIGGCRKQLAQIKE  218
NP_001005677.1[Xenopus_tropicalis]                 RAVEFKVVETDPSPYCIVAPDTVIHCEGEPIKREDEEESLNEVGYDDIGGCRKQLAQIKE  218
                                                   : : * .* .: .   ::   * :   . * :   *. .   : *:**** :::: :::*
```
